# Health professionals’ beliefs and attitudes towards preconception care: A systematic review

**DOI:** 10.1101/2024.09.16.24313739

**Authors:** Cherie Caut, Danielle Schoenaker, Erica McIntyre, Amie Steel

## Abstract

**Background:** Health professionals have previously identified various barriers and factors that would help facilitate preconception care services in healthcare settings. Clinically relevant preconception information and clinical practice guidelines have since been developed to aid health professionals in preconception care delivery. This systematic review aimed to 1) synthesise recent literature (past 8 years) describing health professionals’ beliefs and attitudes towards preconception care services or programmes and 2) determine if the experience of health professionals providing preconception care has changed compared to literature reviews conducted more than 8 years ago.

**Methods:** Five databases were searched between 27/01/2016 and 20/11/2024. Primary quantitative and qualitative research studies were included if they examined health professionals’ beliefs and attitudes towards delivering preconception care services or programmes. Study quality was assessed using the CASP Checklist (qualitative studies) and AXIS tool (quantitative studies). Data synthesis used thematic categorisation adapted from the framework approach.

**Results:** Twenty-seven studies were included (n=11 qualitative, n=14 quantitative, n=2 mixed-methods studies). Methodological quality was generally good for qualitative studies but varied for quantitative studies. The results covered three categories: 1) *addressing preconception care health literacy* (i.e. lack of knowledge, awareness, training and resources), 2) *clinical practicalities of preconception car* e (i.e. need for coordination of care and clarity on role responsibility), and 3) *the role of the patient* (i.e. need for public health education to support patient-led conversations).

**Conclusions:** Little has changed regarding the barriers and facilitators to providing preconception care reported by health professionals. To improve the provision of preconception care, there is a need to co-develop professional and public preconception health education, clinical resources, and a coordinated preconception healthcare service model.

## Introduction

People’s modifiable preconception health (PCH) and health behaviours—such as body composition, lifestyle behaviours, nutrition, environmental exposures, and birth spacing—can affect maternal and child health[1–3]. The need to address such risk factors has led to clinical and public health measures that aim to screen for PCH risks and implement intervention strategies to optimise PCH and the health behaviours of prospective parents before conception, known as preconception care (PCC)[4]. The importance of PCC is highlighted by health policies and strategies produced by a number of countries around the world[5]. It has received dedicated attention from the World Health Organization[4] as a critically important component of healthcare that can impact multiple generations.

While PCC is valuable and important for prospective parents, healthcare providers experience a range of barriers to implementing PCC services or programmes[6, 7]. Previous systematic reviews that included studies conducted more than five years ago have reported health professionals’ (HPs) experiences providing PCC and describe the types of barriers they experience, including but not limited to poor interprofessional communication, insufficient clinical time, funding, clinically relevant information, and public and HP awareness of the benefits of PCC[6, 7]. Conversely, HPs reported that adequate knowledge of PCC, prospective parents discussing their intention to become pregnant or requesting PCC, and clinical PCC resources would enable them to facilitate PCC[7]. To address some of these barriers and promote PCC services, clinician-focussed PCC information and guidelines have been developed[8, 9], and relevant information for implementing PCC programmes in organisations[10]. A recent systematic review, however, identified that existing clinical practice guidelines (CPGs) on PCC supported by high-quality evidence are lacking[11]. Emerging research efforts aim to establish population PCH priorities and co-develop strategies to address them at a healthcare services level[12].

While these guidelines aim to assist HPs providing PCC, an up-to-date understanding of HPs’ views towards PCC delivery is needed to identify if the increasing availability of guidelines has reduced or changed previously reported barriers and if HPs also require other types of support to implement PCC meaningfully. Therefore, this systematic review aimed to 1) synthesise recent literature describing health professionals’ beliefs and attitudes towards preconception care services or programmes and 2) compare these experiences to previous systematic review findings[6, 7].

## Methods

This systematic review is reported following the Preferred Reporting Items for Systematic Reviews and Meta-analyses 2020[13] and was prospectively registered on the International Prospective Register of Systematic Reviews (PROSPERO; CRD42021249386).

### Eligibility criteria

Original primary research (quantitative and qualitative studies) published between 27/01/2016 and 20/11/2024 that sampled HPs (including, but not limited to, general practitioners, midwives, and obstetricians) and examined their beliefs and/or attitudes towards delivering PCC services or programmes were eligible for inclusion.

### Information sources

Keyword and MeSH terms were employed in the databases MEDLINE (OVID), EMBASE (OVID), Maternity and Infant Care (OVID), CINAHL (EBSCO), and PsycINFO (EBSCO), with the following limits: title and abstract, studies in humans, published between 27/01/2016 to 27/01/2022. A second search was conducted for articles published between 28/01/2022 to 20/11/2024. No limits to language were applied. The search strategy is presented in Supplementary File 1.

### Selection process

The search was conducted on 27 January 2022 and 20/11/2024. CC completed electronic database searches and downloaded citations and abstracts into EndNoteX9 citation management software. Duplication screening occurred before citations were exported into Covidence systematic review software[14]. Articles were screened by title and abstract initially by CC, and by DS and AS to establish reliability. Full-text articles were downloaded and screened initially by CC, followed by AS, to establish reliability before final inclusion for review. There were no disagreements between reviewers. References lists of the included studies were searched for additional eligible studies. Article exclusion reasons from the full-text screening stage were recorded.

### Data collection process

CC initially extracted data from eligible studies into a customised form, followed by AS to establish reliability.

### Data items

Data extracted included the study reference, title, aims, type of PCC service, location, population, study design, data collection method(s), sample size, and findings on HP beliefs and attitudes towards PCC.

### Critical appraisal

The Critical Appraisal Skills Programme (CASP) Qualitative Checklist[15] evaluated study reporting and methodological quality. Quantitative studies were assessed via the Appraisal tool for Cross-Sectional Studies (AXIS)[16]. CC critically appraised included studies, and AS assessed 10% of included studies for reliability.

Disagreements were discussed until a consensus was reached. A third reviewer (EM or DS) was invited to adjudicate if unresolved.

### Synthesis methods

A framework approach for applied and policy-relevant research was employed to analyse the data, identify common themes across the studies, and then categorise these findings to determine the key themes[17]. Findings on beliefs and attitudes towards PCC from the included studies were extracted by CC reading and re-reading each paper, and reviewed by AS, DS and EM. Common themes were then identified across the studies and further defined into categories. Each study was assigned as many categories as relevant to their reported findings.

## Results

### Study selection

Searches retrieved 604 articles. After removing duplicates, 376 titles and abstracts were screened (Figure 1). Thirty-seven full-text articles were checked for eligibility; 27 met the inclusion criteria and were included in this review. Reasons for exclusion at full-text screening are presented in Supplementary File 2.

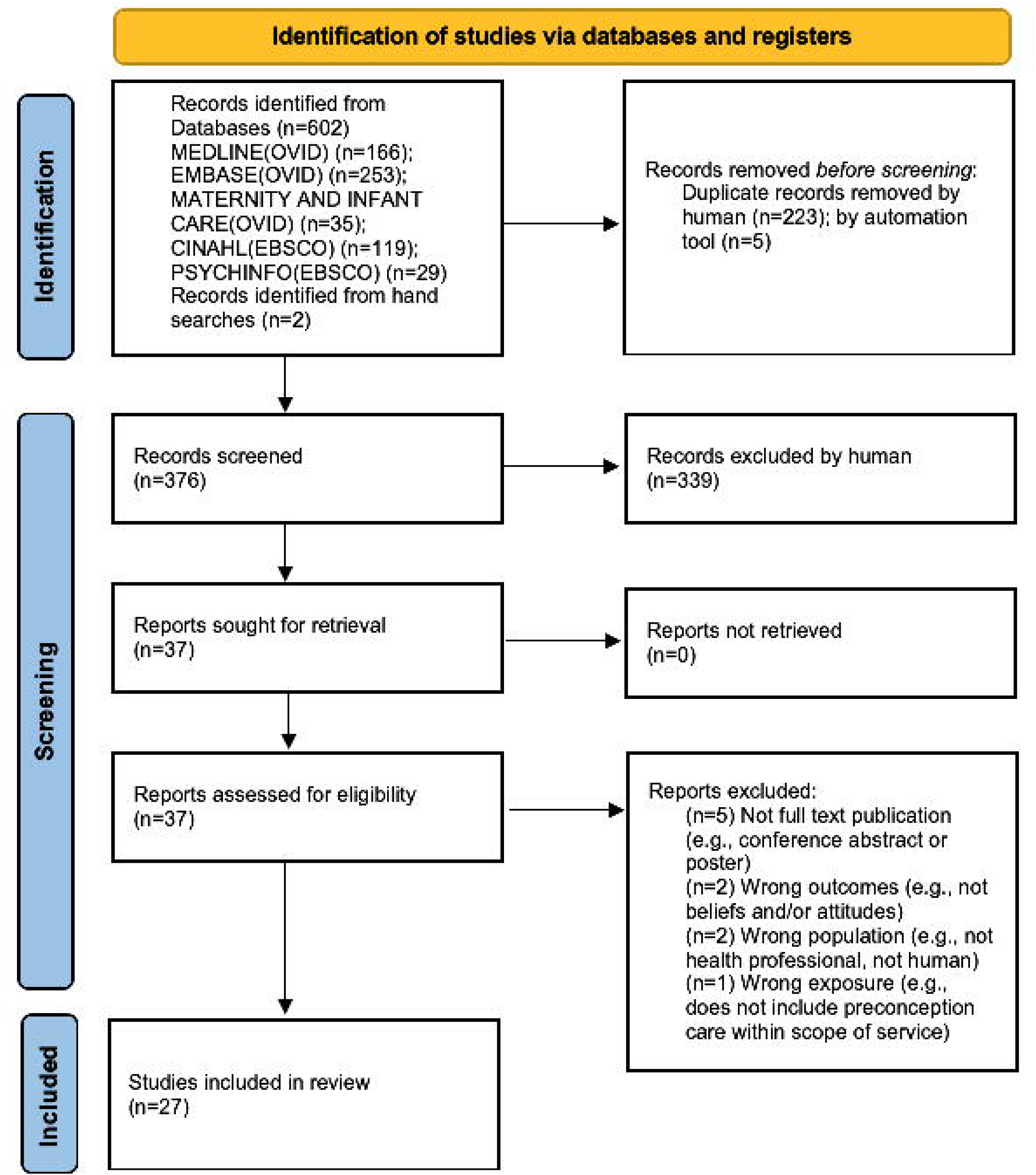

### Study characteristics

All articles (n=27) were published in English between 2017 and 2023 (Table 1). Fourteen studies[18–31] employed a cross-sectional study design and survey method for data collection. Eleven qualitative studies were identified utilising focus groups (n=4)[32–35] or interviews (n=7)[36–42]. Two studies[43, 44] collected data using mixed methods; Devido et al. [43] employed a focus group and survey, and Mass et al. [44] a survey and interactive workshop. The included studies were conducted in Australia (n=6)[21–24, 29, 30]; the United States (US; n=5)[18, 25, 41–43]; the Netherlands (n=4)[31, 33, 37, 44]; United Kingdom (UK) (n=2)[35, 38]; India (n=2)[20, 34]; and one study each in New Zealand[19], Europe[36], Indonesia[32], Canada[26], Malawi[39], South Africa[27], Nigeria[40], and China[28]. A range of HPs were represented, including but not limited to pharmacists, nurses, midwives, general practitioners, obstetricians and gynaecologists. Sample size varied among the qualitative (n=11 to 92), quantitative (n=77 to 992) and mixed-methods (n=48 to 299) studies.

### Critical appraisal of included studies

Quality assessment details are presented in Table 2 and Table 3 for qualitative and quantitative studies, respectively.

Most of the qualitative studies fulfilled all the CASP checklist[15] criteria for assessing methodological quality. One study[39] needed to be more transparent on the appropriateness of the recruitment strategy, consideration of researcher bias and rigour of data analysis. The methodological quality of the quantitative studies was varied[16]. Sample size justification, non-response bias and funding sources or conflicts of interest were the most poorly reported criteria, followed by the use of instruments or measurements that had been trialled, piloted, or published previously.

## Results of syntheses

Three common thematic categories across the findings from the reviewed studies were identified and defined: *addressing preconception care health literacy, clinical practicalities of preconception care*, and *the role of the patient*.

### Addressing preconception care health literacy

Twenty-three studies reported findings related to addressing PCC health literacy[18–26, 28–36, 38, 39, 41–43]. These studies addressed PCH knowledge, clinical training needs and access to resources.

### Preconception health knowledge gaps

Insufficient knowledge about PCH as a barrier to HPs’ PCC practices was commonly reported. For example, an Australian cross-sectional study of GPs (n=110) found their lack of knowledge of PCC guidelines to be one of the most common barriers to their delivery of PCC[24]; this view was shared by 40% of midwives in a second Australian study[21] and 95% of nurses in another Australian study[30], a broader group of HPs from a UK qualitative study.[38] The lack of knowledge as a barrier to PCC provision also applied to GPs (n=304) in Australia providing care to men[23], of whom almost all respondents (90%) indicated that they did not feel confident about their knowledge of the factors affecting male fertility[23]. A cross-sectional study on midwives (n=338) from Australia[29] found that most (85%) rated their overall knowledge about pre and interconception health for women as excellent or above average, although for men or partners, a higher proportion of participants (40%) reported their overall knowledge as below average, poor or none. Participants with more than 11 years of experience were more likely to report above average to excellent knowledge (OR 3.11; 95% CI 1.09, 8.85), although, for men or partners, this association was limited to midwives with more than 21 years of experience (OR 2.20; 95% CI 1.18, 4.11). Another cross-sectional study from Australia[30], on nurses (n=152) found that only one in 20 respondents agreed that they had excellent knowledge about PCC for women and men. When asked to rate their knowledge of the importance of women’s health in the preconception period, over half (54%) reported average to below-average levels. Regarding the importance of men’s health in the preconception period, this increased to two-thirds of respondents (67%) reporting average to below average knowledge. In contrast, a study[28] of HPs in China found that 26.9% of those providing care to women with type 1 diabetes felt they lacked the knowledge to deliver PCC to this population. At the same time, a US study reported a significant positive association between teaching self-efficacy and preconception counselling knowledge among parish nurses (n=48) providing diabetes education to women with diabetes[43]. Only one cross-sectional study sampling dentists and gynaecologists (n=300) from India investigated clinician characteristics associated with PCH knowledge[20]. It was found in logistic regression analyses that older age doctors (Adjusted odds ratio [AOR]=1.055 (95% Confidence interval [CI]: 1.055-1.092, *p*=0.002), the field of specialisation - dentists (AOR=1.635, 95% CI: 1.064-2.511, *p*=0.002), and years of practicing experience > 5 years (AOR=2.254, 95% CI: 1.46-3.45, *p*=0.001) were significantly associated with higher knowledge levels of periodontal health and adverse pregnancy outcomes[20]. One qualitative study from India on healthcare workers (n=45)[34] found that most had some knowledge about preconception care, limited to adolescent health and family planning services. A qualitative study from the UK on pharmacists (n=11)[35] found that lower knowledge scores were related to a lack of training and a lack of confidence in oneself.

### Awareness of preconception risks and guidelines

HPs reported needing to be made aware of PCH risks and of existing CPGs designed to assist them in screening for PCH risks and delivering PCH interventions[19–21, 23, 24, 26, 43]. This issue was highlighted in a study involving dentists and gynaecologists (n=300), where only 79% were aware of the association between periodontal health and preterm low birth weight[20]. In Australia, a study of GPs (n=110) found that only 53% were aware of PCC-CPGs[24]. Similarly, a qualitative study on healthcare providers (n=20) from the US[42] showed a lack of awareness of published PCC guidelines. In contrast, healthcare workers (n=45) in a qualitative study from India were aware that there is no formal PCC program in the country[34]. Even where HPs reported being aware of guidelines, familiarity with the recommendations may pose another barrier, as indicated in an Australian cross-sectional study where fewer HPs (including GPs, obstetricians and gynaecologists, midwives and dietitians) were aware of the recommended dose (38%) or duration (44%) of preconception iodine supplementation[21]. This was despite 71% of those HPs being aware that the National Health and Medical Research Council recommends this intervention[21]. In this Australian study, awareness of the recommendation was positively associated with recommending iodine supplements[21]. In contrast, in a cross-sectional study of GPs (n=200) in New Zealand (NZ), most GPs reported being aware of the risks of obesity in pregnancy, over 50% of these GPs reported practice that was inconsistent with guidelines, and only 12% of these same GPs were aware of the New Zealand Ministry of Health Guidance for Healthy Weight in Pregnancy[19]. A similar finding was found in a Canadian cross-sectional study assessing periconceptional folic acid recommendations of GPs (n=77), where only half knew the correct dose and duration of folic acid for low-risk women[26]. In the Netherlands, most (64.2 %) but not all the respondents in a cross-sectional study on midwives and obstetricians(n=83)[31] believe the scientific proof of the effectiveness of PCC to be sufficient.

### The desire for further training

HPs reported wanting more information and education on PCH risks and interventions[21–23, 33, 39] to improve their confidence in delivering PCC[22, 23, 30]. For example, in a cross-sectional study of Australian health professionals (n=396), HPs (including GPs, obstetricians and gynaecologists, midwives and dietitians) indicated they would be interested in receiving new information relating to iodine[21], and in another Australian cross-sectional study almost all maternal child and family health nurses (MCaFHN) (n=192) agreed that more information and formal education opportunities on the topic would increase their confidence to discuss PCH[22]. Most (87%) nurses (n=152) in another cross-sectional study from Australia[30] indicated that more education would increase their confidence in delivering PCC. Most midwives (n=338) in a cross-sectional study from Australia[29] desired further education on managing pre-existing health conditions (69%) and on optimising reproductive health (66%). Healthcare workers (n=45) suggested the need for program-specific guidelines and training in a qualitative study from India[34]. There was a consensus among pharmacists (n=11) in a qualitative study from the UK[35] that insufficient training opportunities exist. A majority (54.6%) of Australian GPs (n=304) suggested that more information and education about factors that affect male fertility would make them more confident to talk to male patients about fertility[23]. Some studies specifically identified lack of training as the barrier to providing PCC and an increased likelihood of HPs providing PCH information and PCC to their patients if they had received further training[18, 25, 28, 32, 36, 41]. For example, in a cross-sectional study from the US, 15% of GPs (n=443) reported they needed training before implementing PCC[25]. In another cross-sectional study from the US, obstetricians and gynaecologists (n=297) reported offering expanded carrier screening more commonly if they were fellowship-trained (80%) compared to those who were not (70%)[18].

### Access to resources

Ten studies reported findings related to clinician access to resources supporting PCC delivery[21–24, 30, 38, 41, 43]. The types of resources participants suggested include sample meal plans[21], screening guidelines, relevant research findings[21], online tools[22, 30], factsheets, trustworthy websites[22, 23, 30], waiting room posters[23], checklists, brochures[24], expert clinician champions, and medical record prompts[41]. Two other qualitative studies reported structural resource requirements such as PCC policy and procedures used to support them to implement PCC programs into clinical practice settings, as reported in a study of HPs (n=92) from the US[41] and a funding model through third-party reimbursement to improve the delivery of PCC as described by HPs (n=30) in a study from the Netherlands[33].

### Clinical practicalities of preconception care

Twenty studies reported findings related to the clinical practicalities of PCC[20–25, 27–30, 32–35, 37, 39–42, 44].

### Coordination of care

In some studies, HPs reported that a lack of interprofessional referral and care coordination was a barrier to providing PCC[20, 28, 29, 32–34, 37, 41, 44]. One cross-sectional study of dentists and gynaecologists (n=300) from India, for example, found that gynaecologists rarely refer to dentists despite being aware of the link between periodontal health and preterm low birth weight, and only 12% of gynaecologists referred patients to the dentists in the preconception period.[20] Several studies[20, 28, 32, 33] reported HPs describing a desire for an integrated, multi-disciplinary approach to providing PCC services, greater coordination and referral networks between HPs[29, 33, 35, 37, 41, 44] and emphasised the importance of patient follow-up[41]. In a study from the Netherlands, HPs (n=299) suggested that while the responsibility for providing PCC consultations is best suited to primary care, many other HPs involved may act as referrers towards PCC[44].

### Role responsibility

Most HPs in the included studies believed it to be part of their role to provide preconception risk screening, PCH promotion and provision or referral for PCH interventions. These include GPs[23, 24, 30, 44], MCaFHNs[22], midwives[29, 44], nurses[27], pharmacists[35], obstetricians and gynaecologists[42] and specialist physicians[40]. In a qualitative study from the Netherlands, HPs (including midwives, obstetricians and gynaecologists, fertility specialists, GPs, preventive child health care workers, maternity health care providers, physiotherapists, pharmacists and dieticians) (n=30) expressed that the provision of PCC is challenging due to unclear allocation of responsibilities[33]. Although HPs (including midwives, nurses, obstetricians and gynaecologists and GPs) acknowledged that the responsibility to provide PCC consultations lies with all professions[39] in an Australian cross-sectional study, 84% of GPs (n=110) reported that they should be the primary providers of PCC[24]. In a mixed-methods study from the Netherlands, HPs (including nurses, midwives, GPs, physiotherapists, preventive child healthcare professionals, dieticians, policy officers, maternity care assistants, and gynaecologists) (n=299) suggested that the responsibility for providing PCC consultations lies within primary care, mainly GPs (95.6%) and midwives (94.4%)[44]. In this same study, HPs found it significantly more challenging to start a conversation about a wish to conceive than midwives (26.8% versus 20.2%, p=0.006)[44]; they felt less competent to provide preconception information (32.3% versus 15.1%, p=<0.001)[44]. Similar findings were reported in a cross-sectional study from the US (n=443), where most GPs (88%) felt pregnancy intention screening should be routinely included in primary care[25]. In contrast, in a qualitative study from Nigeria, HPs (including nurses and specialist physicians) (n=26) stated that PCC services should be offered at all levels of health care with referral when needed[40]. Specialist physicians from this Nigerian study also identified the relevance of PCC to their practice, stating that those with chronic diseases would benefit more[40]. In a cross-sectional study from Australia on midwives’ (n=338), most (88%) reported that they often encounter health states that could be managed before pregnancy (88%)[29]. Pharmacists (n=11) in a qualitative study from the UK[35] discussed how they are frequently asked for conception advice, particularly by those experiencing difficulties in conceiving or those wanting advice on optimising health to conceive.

### Clinician time

HPs report that one of the barriers to providing effective PCC is an insufficient amount of consultation time[21, 22, 24, 29, 30, 32, 34, 35]. For example, in an Australian cross-sectional study[21], the main reason for not discussing dietary sources of iodine with women was insufficient time[21]. Time constraints were also reported by MCaFHNs (n=192)[22], GPs (n=110)[24], pharmacists[35], and a broader range of HPs (n=32)[32, 34], as the most frequently endorsed barrier to promoting PCH. Several of the MCaFHNs (n=192) suggested that adding a scheduled visit dedicated to interconception health advice to those who want it would be helpful[22].

### The role of the patient

Twelve studies reported findings related to the role of the patient[18, 23, 31, 34, 35, 37–39, 41, 42, 44].

### Community education and health promotion

Four studies indicated that HPs would like to see an increase in public health education that improves their patients’ knowledge of PCH risk factors and the benefits of PCC[18, 37, 44] and perceive the absence of such community awareness as a barrier to PCC[34, 37]. Most (56%) obstetricians and gynaecologists (n=297) believed that offering expanded carrier screening to their patients should be restricted to those diseases important to the couple, and 52% believed that screening should only occur when the clinical significance of each disease being screened for is understood by the couple[18]. In a qualitative study from the Netherlands, HPs (including midwives, GPs, obstetricians and gynaecologists, cardiologists and gastroenterologists) (n=20) stated that barriers affecting the uptake and delivery of PCC included their belief that most future parents lacked awareness of the benefits of PCC[37]. These findings were similar to another qualitative study from India[34], where healthcare workers (n=45) found that preconception was not viewed as a critical phase in the woman’s reproductive cycle, and women would rarely think of eating a balanced diet before pregnancy.

### Patient-led conversations

Despite believing that PCC is a part of their role, HPs also perceive that the onus of responsibility to seek PCH information and PCC is shared with the patient and that improved public education on PCH risks and health behaviours and the benefits of PCC would support this to occur more frequently in the absence of ‘routine’ PCH risk screening and PCH promotion[23, 29–31, 34, 38, 39, 41, 42]. For example, in an Australian cross-sectional study[23], approximately half of GPs’ (n=304) stated that they discuss fertility with male patients ‘opportunistically’ when consulted about a reproductive health matter when a ‘patient specifically asks for advice’ and when consulting with a couple who ‘plan to have children;’ very few said that they raise the subject with men ‘routinely’[23]. In another cross-sectional study (n=152) from Australia[30], 74% of nurses stated they discuss PCC in their practice, although only 13% do so ‘routinely’, and of these, more preconception discussions are held with women than with men. These findings were similar to a qualitative study[41] from the US where the frequency with which HPs (n=92) engaged in conversations about reproductive goals ranged from a ‘routine’ component of each visit to ‘episodically’ or only in response to patients’ ‘question’ or ‘request.’ In a qualitative study from Africa, all the HPs (n=20) felt that women also have a role in demanding PCC services and seeking the services[39]. Conversely, in a cross-sectional study from Australia, midwives (n=338)[29] with a registered nursing qualification were less likely to agree that planning to conceive is a personal decision that should only be discussed when initiated by the woman (OR 0.55; 95% CI 0.35, 0.87).

## Discussion

Findings from this systematic review provide up-to-date insights into the areas that provide opportunities to improve PCC delivery within healthcare settings based on HP’s beliefs and attitudes. These include professional and public health education to increase PCH knowledge and awareness, clinical resources to support HPs providing PCC, and HP referral networks to support effective PCC delivery to prospective parents.

One of the main findings of this review is that, overall, HPs report low PCH literacy yet would like to receive training, suggesting that they may benefit from PCH education. Our findings add to those reported in previous systematic reviews (e.g., HPs wanting more training to improve confidence to provide PCC and lack in knowledge of PCH is a barrier to PCC)[6, 7] indicating little has changed and the PCH education needs of HPs remains to be addressed. There is a high variability of the content and recommendations across existing PCC-CPGs and a need for guidelines that are yet to address all clinical PCC areas[11]. Comprehensive PCC-CPGs that address all evidence-based PCC areas are needed to assist HPs in delivering PCC[11]. However, more than simply developing CPGs to manage the PCH knowledge of HPs is required. As supported by this review’s findings, clinicians often lack awareness of existing guidelines. While some studies have investigated interventions that support the knowledge of HPs and effect changes to practice behaviour[45, 46], further research is needed. Training interventions would benefit from nuanced consideration of the skills, knowledge, time, and funding for targeted HP groups. PCH knowledge may also need to be tiered so foundational training can be included in clinical degrees, with more advanced training available for qualified clinicians. To tailor and co-develop such interventions to specific HPs, further understanding of their knowledge gaps and their current and potential scope of practice concerning PCC is needed.

In this review, some HPs reported believing that the onus of responsibility should be with the patient to seek PCH information and PCC without routine PCH risk screening and PCH promotion. This belief assumes patient awareness of PCH risk factors and the benefits of PCC. With the current level of public health education and understanding of PCH risk factors, this belief may need to be challenged. Similar findings have been reported by HPs in previous systematic reviews (e.g., client’s lack of awareness of PCC, the benefits of PCC and initiation for PCC)[6, 7]. A focus on public education of PCH is also needed to increase the public’s PCH literacy. Adherence to guidelines during preconception is an issue previously described in a systematic review reporting on dietary guideline adherence during preconception and pregnancy[47]. This review also highlighted the importance of acknowledging the influence of demographic and social factors on guideline adherence[47]. Implementation interventions are needed that support knowledge translation and changes to health behaviour through strategies that improve whole population health through the life course (e.g., PCH school education) as well as targeted interventions to those life course phases where becoming pregnant is more likely or possible (e.g., PCH information resources and support tools) or intended (e.g., PCH education and counselling programmes)[48]. As such, Hall and colleagues’ proposed model for PCC integrates education, digital health interventions, campaigns and social media for raising PCH awareness among HPs and the public and includes the individualised and specialised provision of PCC by a range of HPs able to provide clinic-based counselling, motivational interviewing, provision of supplements and interconception interventions[5].

Some HPs report that the need for clinical PCH resources and time are persistent barriers to providing effective PCC. HPs would like access to PCH information resources to support them in delivering PCC. Specifically, HPs identified a range of resources they would find helpful in clinics, from clinician checklists and clinician websites to patient information factsheets and patient websites. Clinicians report that lack of consultation time to provide effective PCC is a barrier, and, in some instances, clinicians would like to be better funded for PCC by receiving third-party reimbursement. Similar findings have been reported by HPs in previous systematic reviews (e.g., a lack of clinical time for PCC and reimbursement for that time, needing physical space or clinically relevant information such as PCC tools checklists and PCC guidelines)[6, 7]. Clinical resources are available, providing organisations and clinicians with information pertinent to implementing PCC programmes[10] and PCC-CPGs (e.g., PCH risk screening checklist or tool and PCC intervention guidelines)[9], yet high-quality guidelines on PCC are lacking[11]. Future focus is needed on assessing the PCH behaviour outcomes associated with healthcare services that implement PCC programmes and when clinicians can access clinically relevant PCH information resources and PCC-CPGs informed by high-quality evidence.

This review also found that many HPs believe they have a role in PCC. However, they also attest that primary care HPs (primarily GPs) should be the leading providers. There are two issues needing further consideration. First, a precedent exists for PCC to be provided by a broader range of HPs, with the opportunity to improve referral networks. Second, these HPs may be under the assumption that the patient’s GP has already provided PCC. These findings are barriers to delivering PCC and were reported by HPs in previous systematic reviews (e.g., a lack of clarity regarding who is responsible for providing PCC[6, 7] and poor communication between HPs)[7]. A PCC model that supports collaboration between HPs and utilisation of all capable HPs[49–51] provides an opportunity to deliver PCC that improves PCH outcomes throughout the community, including at-risk populations and across the life course[12]. Meaningful involvement of all key stakeholders is required to co-design PCC healthcare services that support a coordinated healthcare workforce[50, 51]. Further research is needed to understand the outcomes of an integrated PCC healthcare services delivery model on PCH risks, PCH behaviours and maternal and child health outcomes[5].

### Limitations

This review should be viewed within the context of its limitations. The authors complied with an accepted review methodology[52], although only 10% of the data extraction was verified by a second author. As the data being extracted was not complex and there were no differences identified in the 10% of papers that were checked, the impact of this difference is expected to be minimal. Although the studies in this review represent a wide range of HPs from varied countries or regions, not all HPs who may have a role in PCC were represented in the study populations. The methodological quality of the qualitative studies was good. However, the quantitative study quality was varied. The heterogeneous nature of the study aims and data collection methods precluded the pooling of data for meta-analysis. We should also acknowledge that three studies[19, 33, 37]were included in a previous review[7]. The research question driving our study had a broader focus than the review by Goossens and colleagues and identified an additional 24 papers; however, to avoid overlapping between the two reviews, the findings of the three[19, 33, 37] articles were reported only when other papers in our review shared similar results.

### Conclusions

HPs report insufficient knowledge about PCH, lack PCH training, and want education on PCH risks and interventions to improve their confidence in providing PCC and enhanced access to clinical PCH information resources. One of the barriers they experience to providing effective PCC is an insufficient amount of consultation time. Most HPs believe it is their role to provide PCC, and referral networks could improve the delivery of coordinated preconception interventions to identify and support patients with preconception risks. HPs want increased public health education that improves patient awareness of PCH risks and benefits of PCC. These barriers and facilitators to the provision of PCC reported by HPs in studies conducted in the past five years have remained very similar when compared to previous reviews of earlier studies on the topic. To improve the provision of PCC going forward, there is a need to co-develop professional and public PCH education, PCC clinical resources, and an integrated PCC healthcare service model.

## Supporting information

Table 1. Characteristics of included studies

Table 2. CASP Quality Assessment

Table 3. Axis Quality Assessment

Supplementary Information File 1. Search Strategy

## Data Availability

All data produced in the present work are contained in the manuscript

## List of abbreviations

AXIS: Appraisal tool for Cross-Sectional Studies
CASP: Critical Appraisal Skills Programme
CPG: Clinical practice guideline
GP: General Practitioner
HP: Health professional
MCaFHN: Maternal child and family health nurses
PCC: Preconception Care
PCH: Preconception health
PROSPERO: Prospective Register of Systematic Reviews
UK: United Kingdom
US: United States

## Declarations

### Ethics approval and consent to participate

Not applicable.

### Consent for publication

Not applicable.

### Availability of data and materials

All data generated during this study are included in this published article [and its supplementary information files].

### Authors’ contributions

CC designed the protocol, performed the searches, screening, quality appraisal, data extraction, and data analysis, and drafted, reviewed, and edited the manuscript. DS was the second reviewer for article screening at the title and abstract stage; AS was the second reviewer for article screening at the full-text stage, quality appraisal and data extraction; and AS, DS and EM reviewed and edited the manuscript.

## Acknowledgements

Not applicable.

---

**Supplementary File 1. Search Strategy**

**Supplementary File 2. Full-text articles excluded with reason**

