## Supplementary material for "Health professionals’ beliefs and attitudes towards preconception care: A systematic review": Table 1. Characteristics of included studies

| **Lead author, Year published** | **Location** | | **Study aim** | **Component of PCC studied** | **Study design** | **Data collection method** | **Target population** | **Sample size** | **Themes** | | |
| --- | --- | --- | --- | --- | --- | --- | --- | --- | --- | --- | --- |
|  |  | |  |  |  |  |  |  | I | II | III |
| Bradfield 2023 | Australia | | Explore midwives’ knowledge, perspectives and learning needs and barriers and enablers to delivering preconception and interconception care. | General preconception and interconception care provision | Cross-sectional study | Survey | Midwives | N=338 | • | • | • |
| Briggs 2018 | United States | | Evaluate physician’s utilisation and attitudes towards expanded carrier screening. | Genetic screening and counselling | Cross-sectional study | Survey | OBGYNs | N=297 | • |  | • |
| Chutke 2022 | India | | Identify barriers and suggestions for framing appropriate strategies for implementing preconception care through primary health centres. | General preconception care provision | Qualitative study | Focus group | Healthcare workers: Nurse, Midwife, Multi purpose worker | N=45 | • | • | • |
| Devido, 2017 | United States | | Explore the role and experiences of the parish nurse in providing diabetes education and preconception counselling to women with diabetes. | Diabetes education | Mixed methods | Focus group  Survey | Nurses | N=48 | • |  |  |
| Dorney 2022 | Australia | | Understand primary healthcare nurses knowledge, practice and attitudes to preconception care, | General preconception care provision | Cross-sectional study | Survey | Primary healthcare nurses | N=152 | • | • | • |
| Fieldwick, 2017 | New Zealand | | Explore the knowledge and practice of New Zealand general practitioners regarding preconception and gestational weight management. | Gestational weight management | Cross-sectional study | Survey | General practitioners | N=200 | • |  |  |
| Ganganna, 2017 | India | | Evaluate the knowledge, attitude, and practices of dentists‚ and gynaecologists’‚ concerning their role in educating patients toward maintaining good oral health. | Oral hygiene and periodontal disease | Cross-sectional study | Survey | Dentists and Gynaecologists | N=300 | • | • |  |
| Guess 2017 | Australia | | Assess knowledge and practices of Australian healthcare providers in relation to the National Health and Medical Research Council‚ iodine supplement recommendation. | Iodine supplementation | Cross-sectional study | Survey | Midwives; General practitioners; Obstetricians; Gynaecologists; Dietitians | N=396 | • | • |  |
| Hammarberg, 2019 | Australia | | Assess Maternal, Child and Family Health Nurses attitudes towards preconception health promotion, whether and under what circumstances they talk to their families about this, and what might help them start a conversation about preconception health. | General preconception health promotion | Cross-sectional study | Survey | Nurses; Midwives | N=192 | • | • |  |
| Hogg 2019 | Australia | | Men’s preconception health care in Australian general practice: GPs’ knowledge, attitudes, and behaviours | Men’s preconception health care | Cross-sectional study | Survey | General practitioners | N=304 | • | • | • |
| Janssens, 2017 | Europe | | Explore clinical and molecular geneticists' views on the implementation of expanded carrier screening in the clinical setting. | Genetic screening and counselling | Qualitative study | Interview | Geneticists | N=16 | • |  |  |
| Kizirian, 2019 | Australia | | Explore the understanding and provision of preconception care by general practitioners within the Sydney Local Health District. | General preconception care provision | Cross-sectional study | Survey | General practitioners | N=110 | • | • |  |
| Kurniawati, 2021 | Jakarta, Indonesia | | Identifying the nature of preconception care delivered by healthcare practitioners toward prospective brides and grooms at primary healthcare in Jakarta. | General preconception care provision | Qualitative study | Focus group | Nurses; Midwives; General practitioners; Psychologists | N=32 | • | • |  |
| M'Hamdi, 2017 | Netherlands | | Examine health care professionals‚ views of their role and responsibilities in providing preconception care and identify barriers that affect the delivery and uptake of preconception care. | General preconception care provision | Qualitative study | Interview | Midwives; General practitioners; Obstetricians; Cardiologist; Gastroenterologist | N=20 |  | • | • |
| Maas, 2022 | Netherlands | | Explore healthcare providers views on improving preconception care in their region. | General preconception care provision | Mixed methods | Survey and interactive workshop | Nurses; Midwives; General practitioners; Physiotherapist; Preventive child healthcare professional; Dietician, Policy officer; Maternity care assistant; Gynaecologist | N=299 |  | • | • |
| Manze, 2020 | United States | | Assess factors associated with routine pregnancy intention screening by primary care physicians and their support for such an initiative. | Pregnancy intention screening and counselling | Cross-sectional study | Survey | General practitioners | N=443 | • | • |  |
| Mida, 2021 | Canada | | Assess the knowledge, attitude and practice of physicians regarding periconceptional folic acid recommendations, intakes, and health related outcomes for women at low risk of a neural tube defect affected pregnancy. | Multivitamin and folic acid supplementation | Cross-sectional study | Survey | Family physicians | N=77 | • |  |  |
| Molinaro, 2021 | United Kingdom | | Explore:1) how clinicians providing care to patients from preconception to the first 2 years of life and treating chronic diseases like to learn about DOHaD; 2) what factors influence counselling on developmental programming to patients; and 3) how knowledge translation about DOHaD can be enhanced in reproductive health care practice. | General preconception health promotion | Qualitative study | Interview | Obstetrician/gynaecologists; family physicians/General Practitioners; Midwives; Endocrinologists; Internal medicine generalists; Maternal fetal medicine specialists; and Paediatricians. | N=23 | • |  | • |
| Munthali, 2021 | Malawi, Africa | | Examine knowledge and perceptions of preconception care among health workers and women of reproductive age. | General preconception care provision | Qualitative study | Interview | Nurses; Midwives; General practitioners; Gynaecologist; Medical doctor; Clinical officer; Nurse-midwife officer; Nurse-midwife technician | N=20 | • | • | • |
| Nacer 2022 | United States | | Examine the factors influencing behaviour of health care providers around preconception care outpatient clinical settings in the United States. | General preconception care provision | Qualitative study | Interview | Healthcare Providers: Family Medicine Physicians,Obstetricians/Gynecologists, Nurse Practitioners, Nurse, Midwife | N=20 | • | • | • |
| Ojifinni, 2022 | Nigeria | | Explore perceptions about preconception care services among health care providers. | General preconception care provision | Qualitative study | Interview | Nurses; Specialists physicians | N=26 |  | • |  |
| Poels, 2017 | Netherlands | | Identify bottlenecks and solutions for the delivery of preconception care from a healthcare providers perspective in a local community setting in the Netherlands. | General preconception care provision | Qualitative study | Focus group | General practitioners; Midwives; Gynaecologists; Physiotherapists; Fertility specialists; Pharmacists; Preventive child healthcare; Maternity care; Dietician; Municipal policy officer; Patient advocacy | N=30 | • | • |  |
| Scheele 2023 | Netherlands | | Evaluate the current practice of preconception care in the Netherlands and the perceptions of birth care professionals concerning preconception care. | General preconception care provision | Cross-sectional study | Survey | Midwives and Obstetricians | N=83 | • |  | • |
| Silverio 2023 | United Kingdom | | Explore community pharmacists’ practices and attitudes towards the provision of healthcare advice regarding preconception and pregnancy. | General preconception care provision | Qualitative study | Focus group | Pharmacists | N=11 | • | • |  |
| Simone, 2018 | United States | | Describe current models of preconception care and explore factors influencing services. | General preconception care provision | Qualitative study | Interview | Nurses; General practitioners; HIV healthcare providers including Physicians; Nurse practitioners; Physician assistants; Nurses; Social worker | N=92 | • | • | • |
| Ukoha, 2019 | South Africa | | Describe the preconception health care nursing student‚ knowledge of and attitude towards the provision of preconception care. | General preconception care provision | Cross-sectional study | Survey | Nurses | N= 138 |  | • |  |
| Wang, 2021 | China | | Investigate the current practice and perspectives of healthcare providers regarding preconception care for women with type 1 diabetes in China. | Diabetes education | Cross-sectional study | Survey | Nurses; Midwives; Endocrinologists, Diabetes nurses, Gynaecologists, Obstetricians | N=992 | • | • |  |
|  | | **Themes: I: Addressing preconception care health literacy II: Clinical practicalities of preconception care III: The role of the patient** | | | | | | | | | |
