## Supplementary material for "Health professionals’ beliefs and attitudes towards preconception care: A systematic review": Table 2. CASP Quality Assessment

|  | **Chutke 2022** | **Devido 2017** | **Janssens 2017** | **Kurniawati 2021** | **M'Hamdi 2017** | **Maas 2022** | **Molinaro 2021** | **Munthali 2021** | **Nacev 2022** | **Ojifinni 2022** | **Poels 2017** | **Silverio 2023** | **Simone 2018** |
| --- | --- | --- | --- | --- | --- | --- | --- | --- | --- | --- | --- | --- | --- |
| 1. Was there a clear statement of the aims of the research? | Yes | Yes | Yes | Yes | Yes | Yes | Yes | Yes | Yes | Yes | Yes | Yes | Yes |
| 2. Is a qualitative methodology appropriate? | Yes | Yes | Yes | Yes | Yes | Yes | Yes | Yes | Yes | Yes | Yes | Yes | Yes |
| 3. Was the research design appropriate to address the aims of the research? | Yes | Yes | Yes | Yes | Yes | Yes | Yes | Yes | Yes | Yes | Yes | Yes | Yes |
| 4. Was the recruitment strategy appropriate to the aims of the research? | Yes | Yes | Yes | Yes | Yes | Yes | Yes | Can’t tell | Yes | Yes | Yes | Yes | Yes |
| 5. Was the data collected in a way that addressed the research issue? | Yes | Yes | Yes | Yes | Yes | Yes | Yes | Yes | Yes | Yes | Yes | Yes | Yes |
| 6. Has the relationship between researcher and participants been adequately considered? | Yes | Yes | Yes | Yes | Yes | Yes | Yes | Can’t tell | Yes | Yes | Yes | Yes | Yes |
| 7. Have ethical issues been taken into consideration? | Yes | Yes | Yes | Yes | Yes | Yes | Yes | Yes | Can’t tell | Yes | Yes | Yes | Yes |
| 8. Was the data analysis sufficiently rigorous? | Yes | Yes | Yes | Yes | Yes | Yes | Yes | Can’t tell | Yes | Yes | Yes | Yes | Yes |
| 9. Is there a clear statement of findings? | Yes | Yes | Yes | Yes | Yes | Yes | Yes | Yes | Yes | Yes | Yes | Yes | Yes |
| 10. How valuable is the research? | Yes | Yes | Yes | Yes | Yes | Yes | Yes | Yes | Yes | Yes | Yes | Yes | Yes |
