## Supplementary material for "Health professionals’ beliefs and attitudes towards preconception care: A systematic review": Table 3. Axis Quality Assessment

|  | Bradfield 2023 | Briggs 2018 | Devido 2017 | Dorney 2022 | Fieldwick 2017 | Ganganna 2017 | Guess 2017 | Hammarberg 2019 | Hogg 2019 | Kizirian 2019 | Maas 2022 | Manze 2020 | Mida 2021 | Scheele 2023 | Ukoha 2019 | Wang 2021 |
| --- | --- | --- | --- | --- | --- | --- | --- | --- | --- | --- | --- | --- | --- | --- | --- | --- |
| 1. Were the aims/objectives of the study clear? | Yes | Yes | Yes | Yes | Yes | Yes | Yes | Yes | Yes | Yes | Yes | Yes | Yes | Yes | Yes | Yes |
| 2. Was the study design appropriate for the stated aim(s)? | Yes | Yes | Yes | Yes | Yes | Yes | Yes | Yes | Yes | Yes | Yes | Yes | Yes | Yes | Yes | Yes |
| 3. Was the sample size justified? | Yes | No | No | No | Yes | No | No | No | No | No | No | No | No | No | Yes | Yes |
| 4. Was the target/reference population clearly defined? (Is it clear who the research was about?) | Yes | Yes | Yes | Yes | Yes | Yes | Yes | Yes | Yes | Yes | Yes | Yes | Yes | Yes | Yes | Yes |
| 5. Was the sample frame taken from an appropriate population base so that it closely represented the target/reference population under investigation? | Yes | Yes | Yes | Yes | Yes | Yes | Yes | Yes | Yes | Yes | Yes | Yes | Yes | Yes | Yes | Yes |
| 6. Was the selection process likely to select subjects/participants that were representative of the target/reference population under investigation? | Yes | Yes | Yes | Yes | Yes | Yes | Yes | Yes | Yes | Yes | Yes | Yes | Yes | Yes | Yes | Yes |
| 7. Were measures undertaken to address and categorise non-responders? | Yes | Yes | No | Yes | Yes | Yes | Yes | Yes | Yes | Do not know | Yes | Yes | No | Yes | Yes | Yes |
| 8. Were the risk factor and outcome variables measured appropriate to the aims of the study? | Yes | Yes | Yes | Yes | Yes | Yes | Yes | Yes | Yes | Yes | Yes | Yes | Yes | Yes | Yes | Yes |
| 9. Were the risk factor and outcome variables measured correctly using instruments/ measurements that had been trialled, piloted or published previously? | Yes | No | Yes | Yes | Yes | Yes | Yes | Yes | Yes | Yes | No | Yes | No | Yes | Yes | Yes |
| 10. Is it clear what was used to determined statistical significance and/or precision estimates? (eg, p values, CIs) | Yes | Yes | Yes | Yes | Yes | Yes | Yes | Yes | Yes | Yes | Yes | Yes | Yes | Yes | Yes | Yes |
| 11. Were the methods (including statistical methods) sufficiently described to enable them to be repeated? | Yes | Yes | Yes | Yes | Yes | Yes | Yes | Yes | Yes | Do not know | Yes | Yes | Yes | Yes | Yes | Yes |
| 12. Were the basic data adequately described? | Yes | Yes | Yes | Yes | Yes | Yes | Yes | Yes | Yes | Yes | Yes | Yes | Yes | Yes | Yes | Yes |
| 13. Does the response rate raise concerns about non-response bias? | No | Do not know | Do not know | No | Yes | No | Do not know | Yes | No | Yes | Do not know | No | Do not know | No | No | No |
| 14. If appropriate, was information about non-responders described? | Yes | Yes | No | Yes | Yes | Yes | Yes | No | Yes | Yes | Yes | Yes | No | Yes | Yes | Yes |
| 15. Were the results internally consistent? | Yes | Yes | Yes | Yes | Yes | Yes | Yes | Do not know | Yes | Do not know | Yes | Yes | Yes | Yes | Yes | Yes |
| 16. Were the results for the analyses described in the methods, presented? | Yes | Yes | Yes | Yes | Yes | Yes | Yes | Yes | Yes | Yes | Yes | Yes | Yes | Yes | Yes | Yes |
| 17. Were the authors‚ discussions and conclusions justified by the results? | Yes | Yes | Yes | Yes | Yes | Yes | Yes | Yes | Yes | Yes | Yes | Yes | Yes | Yes | Yes | Yes |
| 18. Were the limitations of the study discussed? | Yes | Yes | Yes | Yes | Yes | No | Yes | Yes | Yes | Yes | Yes | Yes | Yes | Yes | Yes | Yes |
| 19. Were there any funding sources or conflicts of interest that may affect the authors‚ interpretation of the results? | No | Yes | Yes | No | Yes | No | No | No | No | Do not know | No | Do not know | No | No | No | No |
| 20. Was ethical approval or consent of participants attained? | Yes | Yes | Do not know | Yes | Yes | Yes | Yes | Yes | Yes | Yes | Yes | Yes | Yes | Yes | Yes | Yes |
