## Supplementary Information File 1. Search Strategy for "Health professionals’ beliefs and attitudes towards preconception care: A systematic review"

### Supplementary File 1. Search strategy

#### Search terms

|  | Boolean operator ‘OR’ |  | Boolean operator ‘OR’ |  | Boolean operator ‘OR’ |
| --- | --- | --- | --- | --- | --- |
| MeSH terms |  |  |  |  |  |
|  | Preconception Care | AND | General Practitioners  Health Personnel  Midwifery  Nurses  Physicians | AND | Attitude  Attitude of Health Personnel |
| Key words | before pregnancy  interconception  inter-conception  interpregnancy  inter-pregnancy  preconception  pre-conception  preconception care  periconceptional  peri-conceptional  pre-pregnancy  prepregnancy |  | general practitioner  gynaecologist  gynecologist  health personnel  healthcare provider  health care provider  healthcare professional  health care professional  midwife*  nurse*  obstetrician  physician* |  | acceptability  attitude  belief*  experience*  perception*  attitude |
| MeSH terms and key words will be combined with Boolean operator ‘OR’ and each column combined with the Boolean operator ‘AND’ | | | | | |

#### Search strategy

MEDLINE (OVID), EMBASE (OVID), Maternity and Infant Care (OVID), humans, articles published in the last 8 years. MeSH terms and keywords will be searched by title and abstract. MeSH terms containing relevant trees will be exploded.

SEARCH 1: ((exp Preconception Care.sh OR (before pregnancy OR interconception OR inter-conception OR interpregnancy OR inter-pregnancy OR preconception OR pre-conception OR preconception care OR periconceptional OR peri-conceptional OR pre-pregnancy OR prepregnancy).tw.)

AND

SEARCH 2: (exp (General Practitioners OR Health Personnel OR Midwifery OR Nurses OR Physicians).sh OR (general practitioner OR gynaecologist OR gynecologist OR healthcare provider OR health care provider OR healthcare professional OR health care professional OR midwife* OR nurse* OR obstetrician OR physician*).tw.))

SEARCH 3: (exp (Attitude OR Attitude of Health Personnel).sh OR (acceptability OR attitude OR belief* OR experience* OR perception*).tw.))

SEARCH 4: 1 AND 2 AND 3

CINAHL (EBSCO), humans, articles published in the last 8 years. Keywords will be searched by title and abstract. Subject headings containing relevant trees will be exploded. MH CINAHL Subject Heading, MH+ Exploded CINAHL Subject Heading, TI Title, AB Abstract.

1. Preconception Care MH+
2. “before pregnancy” TI OR AB
3. interconception TI OR AB
4. inter-conception TI OR AB
5. interpregnancy TI OR AB
6. inter-pregnancy TI OR AB
7. preconception TI OR AB
8. pre-conception TI OR AB
9. “preconception care” TI OR AB
10. periconceptional TI OR AB
11. peri-conceptional TI OR AB
12. pre-pregnancy TI OR AB
13. prepregnancy TI OR AB
14. 1 OR 2 OR 3 OR 4 OR 5 OR 6 OR 7 OR 8 OR 9 OR 10 OR 11 OR 12 OR 13 OR 14
15. General Practitioners MH+
16. Health Personnel MH+
17. Midwifery MH+
18. Nurses MH+
19. Physicians MH+
20. “general practitioner” TI OR AB
21. gynaecologist TI OR AB
22. gynecologist TI OR AB
23. “healthcare provider” TI OR AB
24. “health care provider” TI OR AB
25. “healthcare professional” TI OR AB
26. “health care professional” TI OR AB
27. midwife* TI OR AB
28. nurse* TI OR AB
29. obstetrician TI OR AB
30. physician* TI OR AB
31. 15 OR 16 OR 17 OR 18 OR 19 OR 20 OR 21 OR 22 OR 23 OR 24 OR 25 OR 26 OR 27 OR 28 OR 29 OR 30
32. Attitude MH+
33. Attitude of Health Personnel MH+
34. acceptability TI OR AB
35. attitude TI OR AB
36. belief* TI OR AB
37. experience* TI OR AB
38. perception* TI OR AB
39. 32 OR 33 OR 34 OR 35 OR 36 OR 37 OR 38
40. 14 AND 31 AND 39

PsycINFO (EBSCO), humans, articles published in the last 8 years. Keywords will be searched by title and abstract. TI Title, AB Abstract, MA MeSH Subject Heading

1. “Preconception Care” MA
2. “before pregnancy” TI OR AB
3. “interconception” TI OR AB
4. “inter-conception” TI OR AB
5. “interpregnancy” TI OR AB
6. “inter-pregnancy” TI OR AB
7. “preconception” TI OR AB
8. “pre-conception” TI OR AB
9. “preconception care” TI OR AB
10. “periconceptional” TI OR AB
11. “peri-conceptional” TI OR AB
12. “pre-pregnancy” TI OR AB
13. “prepregnancy” TI OR AB
14. 1 OR 2 OR 3 OR 4 OR 5 OR 6 OR 7 OR 8 OR 9 OR 10 OR 11 OR 12 OR 13 OR 14
15. “General Practitioners” MA
16. “Health Personnel” MA
17. “Midwifery” MA
18. “Nurses” MA
19. “Physicians” MA
20. “general practitioner” TI OR AB
21. “gynaecologist” TI OR AB
22. “gynecologist” TI OR AB
23. “healthcare provider” TI OR AB
24. “health care provider” TI OR AB
25. “healthcare professional” TI OR AB
26. “health care professional” TI OR AB
27. “midwife” TI OR AB
28. “nurse” TI OR AB
29. “obstetrician” TI OR AB
30. “physician” TI OR AB
31. 15 OR 16 OR 17 OR 18 OR 19 OR 20 OR 21 OR 22 OR 23 OR 24 OR 25 OR 26 OR 27 OR 28 OR 29 OR 30
32. “Attitude” MA
33. “Attitude of Health Personnel” MA
34. “Acceptability” TI OR AB
35. “attitude” TI OR AB
36. “belief” TI OR AB
37. “experience” TI OR AB
38. “perception” TI OR AB
39. 32 OR 33 OR 34 OR 35 OR 36 OR 37 OR 38
40. 14 AND 31 AND 39
